# Understanding SARSCOV-2 propagation, impacting factors to derive possible scenarios and simulations

**DOI:** 10.1101/2020.09.07.20190066

**Authors:** Lewis Mehl-Madrona, François Bricaire, Adrian Cuyugan, Jovan Barac, Asadullah Parvaiz, Ali Bin Jamil, Sajid Iqbal, Ryan Vally, Meryem Koliali, Mohamed Karim Sellier

**Affiliations:** Bangor (Northern Light Family Medicine Residency) and Orono, Maine (University of Maine and Coyote Institute); Université de la Sorbonne – Paris, France; Peritus – Manila, The Philippines; Peritus – Sydney, Australia; Aspire Technologies – Copenhagen, Denmark; Codehoppers – Lahore, Pakistan; BCB – Paris, France; The Innate – London, UK

**Keywords:** Covid-19 SARSCOV-2 propagation mortality heating, ventilation, air conditioning (HVAC), vitamin D obesity masking policies crowdedness

## Abstract

**Objectives:** We aimed to analyze factors impacting the Covid-19 epidemic on a macro level, comparing multiple countries across the world, and verifying the occurrence at a micro level through cluster analysis.

**Design:** Statistical analysis of large datasets.

**Methods:** We used publicly available large world datasets (1-11). Data was transformed to fit parametric distributions prior to statistical analyses, which were performed with Student’s t-test, linear regression and post-hoc tests. Especially for ordinary least squares regression, natural logarithmic transformations were done to remediate normality violations in the standardized residuals.

**Results:** The severity of the epidemic was most strongly related to exposure to ultraviolet light and extrapolated levels of vitamin D and to the health of the population, especially with regards to obesity. We found no county with an obesity level < 8% with a severe epidemic. We also found that countries where the population benefited from sun exposure or vitamin D supplementation and spent time outside fared well. Factors related to increased propagation of the virus included the use of heating ventilation and air conditioning (HVAC), population density, poorly aerated gatherings, relative humidity, timely policies of closing clustering places until aeration was improved, and daily amount of ridership on public transportation, especially subways. Population lockdowns, masks, and blood type did not provide much explanatory power. The excess mortality observed is within the ranges of severe past influenza epidemics of 2016/2017 or 1999/2000.

**Conclusions:** Our study suggested that prevention measures should be directed to improving aeration systems, enhancing diets and exercise, and ensuring adequate levels of vitamin D. Further research on masking is indicated as our study could not separate policies from how well they were actually followed.

**Funding:** This research received no specific grant from any funding agency in the public, commercial or not-for-profit sectors’

**Strengths and Limitations of the Study:**

- The Study examines large datasets across countries to look for macrotrends in management of the Covid-19 outbreak.
- The Study cannot necessarily establish causation but rather correlation.
- The Study raises some novel possibilities for further studies in relation to country-wide and individual-level susceptibility to Covid-19 and to other epidemics in general.
- The Study raises questions about some political policies based upon country-level comparisons and suggests some areas for exploration of prevention policies.

## Introduction

The data needed to accurately track the transmission and impact of COVID-19 have been hard to collect. The available data do not provide the full picture of the epidemic, are not standardized among countries, and are not always standardized within regions of the same country. Understandably, most governments (and modelers) have focused their efforts on in-country tracking and predictions, making international comparisons difficult. Data are defined and collected differently from country to country, from period to period (and sometimes even within a country at the same period).

For an epidemic to be understood, we must understand environmental factors (which can include hours of daylight, temperature, humidity, ease of absorption of vitamin D, diet, and the presence of other pathogens, host factors (lifestyle, past and present exposure to other pathogens, obesity, co-morbid conditions, age, hygiene, mask wearing, and treatments), transmission factors (aerosol, hands, droplets, intermediary agent), and context (urban or rural, population density, types of heating and cooling, types of transportation, and gatherings, shared toilets) for example. The examination of large datasets and the comparisons of countries with each other can provide important clues regarding the behavior of the virus and this epidemic and can give clues regarding optimum management strategies. Variables work together so that their influence may sometimes only be seen in large datasets.

Epidemic clusters have been identified involving closed places with people gathering with little air circulation such as transportation centers, places of worship, slaughterhouses, companies, hospitals, care facilities, prisons, jails, and police stations. On a more general level, major cities including London, New York, Paris, Madrid, and Milan, with factories, air-conditioned offices, dense public transportation, and regular crowed social and religious events offer higher superspreading opportunities than medium cities or rural areas where little superspreading happened.

These observations guided our efforts to analyze some large datasets to determine what factors would emerge as most important for propagation and host resistance.

## Methods

We formed a multi-disciplinary team that included an infectious diseases physician, data scientists, and software developers. We continuously challenged our projections with reality and numbers from countries around the world, allowing us to refine our models and better understand the progression of the epidemic. All our predictions and findings were discussed and validated from a clinical viewpoint. We used publicly available large datasets [1-11]

### Data Preparation and Analysis

Data was transformed to fit parametric distributions prior to statistical analyses, which were performed with Student’s t-test, linear regression and post-hoc tests. Especially for ordinary least squares regression, natural logarithmic transformations were done to remediate normality violations in the standardized residuals. Interpretation on the final model of the regression and analysis of variance were adjusted because of logarithmic transformations. For correlational studies, whether continuous or count data were used, both Pearson’s and Spearman’s rank correlational tests were done according to the data used. Normality assumptions were not made as we used non-parametric correlation tests to overcome nonlinearity. Most of these normality violations related to bimodal distribution, so running Pearson’s correlation tests provided a complete description of the association [12]. Also, power in significance testing were also done to ensure that for some data points that have very minimal number of samples, this was considered that the interpretation of significance is practically taken into consideration. For cross-correlation of stochastic processes proper diagnostics were done to ensure the absence of autocorrelation processes that could signal the delayed copy of itself from its own function of lag. Some weakpoints of the analysis that a limited area (i.e. UV Index, ozone and average daily ridership) were correlated to the national level counts or vice-versa with case counts, death counts, density GDP, etc with daily ridership. This is due to limited public data available. All significance levels are set at 0.05, otherwise specifically stated.

### Excess mortality

was assessed using Euromomo charts, in which data points above the red line represent increases in excess mortality. Euromomo charts provide z-scores for excess deaths. This was cross-referenced with the Oxford dataset. The Oxford COVID-19 Government Response Tracker told us when each country imposed lockdowns and other restrictions. Spain imposed a lockdown on 14 March when they had 6300+ cases and almost 200 deaths. Italy imposed a targeted stay at home order on 23 February and a general population lockdown on 10 March when they had 10k+ cases and 630+ deaths. France imposed a lockdown on 17 March when they had 7.6k+ cases and 140+ deaths. The UK imposed a general population lockdown on 23 March when they had 12k+ cases and 360+ deaths. Greece imposed a lockdown on 23 March when they had 650+ cases. Malta imposed restrictions on gatherings on 10 March when they had 5 cases. Portugal imposed a lockdown on 19 March when they had almost 800 cases. Belgium imposed lockdown on 13 March when they had 559 cases and 3 deaths. Restrictions on large gatherings and public places were obtained from the Oxford data and from the Wiki France COVID timeline. Japan imposed a stay at home order on 8 April when they had 4400+ cases and almost 100 deaths (Wikipedia history of events). Tajikistan imposed a lockdown on 9 May when they had 610+ cases and 20 deaths, while Kazakstan imposed a strict lockdown on 19 March when they had 44 cases and no deaths yet. [April 1st (Week 14) on Euromomo was the highest peak in excess deaths. The Centre for the Mathematical Modeling of Infectious Diseases published estimated R_0_ figures and rates of growth and doubling time per country (https://dataverse.harvard.edu/dataverse/covid-rt).

### Environmental relationships

were tested using Pearson correlation analysis. The plots and values can be downloaded from https://peritusservices-my.sharepoint.com/:p:/g/personal/adrian_peritus-services_com/EeQWxQGX-s5PjfC5cbf2Bw8BHxFs9Njywh06FULKsyFyEA?e=XkPYn4. The relative humidity of cities was taken and aggregated into monthly average and quarterly average and then correlated with the national level counts of cases of deaths per country. This was found to be significantly correlated at 0.10 alpha (see link for the plot). UV Index and Ozone levels were taken from the TEMIS satellite dataset, which was cross correlated with the daily national level counts of cases and deaths per country (see https://public.tableau.com/profile/foxyreign#!/vizhome/UVIndexandOzone/Cross-Correlation). The cross-correlation dashboard based on selected European cities can be viewed at https://public.tableau.com/profile/foxyreign#!/vizhome/UVIndexandOzone/Cross-Correlation. This data does not adjust for average levels of melanin in each country. UV Index and Ozone Cross-Correlation data are found at https://public.tableau.com/profile/foxyreign#!/vizhome/UVIndexandOzone/Cross-Correlation.

We used Wikipedia pages of selected countries to obtain GINI Index, GDP per Capita, Density, Population count and Average Daily Ridership. We used Accuweather, Weather.com, and Weatherspark.com to obtain data on sunshine and relative humidity as monthly averages. Analysis was done with pair-wise Pearson product-moment correlations. We used 18 European cities, including Paris, Herault, Bouches du Rhone, Loire Atlantique, Nord, Brussels Capital Region, Community of Madrid, Barcelona, MilanRome, Porto Metro Area (North), Lisbon Metro Area, Copenhagen Urban, Stockholm County, Rio de Janeiro State, Sao Paolo State, Miami-Dade County, and New York City. Case counts, death counts, cases per capita and deaths per capita exhibited skewed distributions. Non-parametric methods were used.

Time-series with lag regression was used to compute the **mean incubation period** for change in percentage of deaths and not the absolute count of deaths. The screenshot of the model summaries are available at https://peritusservices-my.sharepoint.com/:p:/g/personal/adrian_peritus-services_com/EQhDar3AuGZAhRmUvvrKmyABpfHbSGJ8xdkMs7vnKg0K7Q?e=1K3KHy. Pre-selection of the lag to be included in the lag regression was done using the cross-correlation but the time-series did not undergo pre-whitening nor was it adjusted for seasonality.

### Data sources included the

UV station data based on TEMIS satellite ozone data, available at https://www.temis.nl/uvradiation/UVarchive/stations_uv.php (Cloud-free erythemal UV Index and local solar noon ozone values, date range from 1 Mar to 25 May 2020), COVID-19 Coronavirus Pandemic, Worldometers, date range from 1 Mar to 25 May 2020, and COVID-19 Community Mobility Reports, Google, date range from 1 Mar to 25 May 2020 - https://www.gstatic.com/covid19/mobility/Global_Mobility_Report.csv. Selected cities were joined that matched between Google Mobility and TEMIS Satellite datasets. There were 7 cities included Venice, Paris, Madrid, Copenhagen, Bern, Stockholm, and Lisbon. The daily death change percentage has extreme values so any death change percentage above 200% was replaced, setting the limit to 200% (i.e. the maximum was 3,600% Sweden from 2 to 80+ deaths in just two days!). We ran the 1^st^ iteration of the model using the computed for cross-correlation of available data and picked out the top 3 correlation coefficient values under 0.05 significance:

∘ UV Index – lag +7 and +11
∘ Transit – lag +11, +17, +24
∘ Residential – lag 10, 17, 24

Then we ran the 2^nd^ iteration of the model using the computed for cross-correlation of available data and picked out the highest correlation coefficient values under 0.05 significance:

∘ UV Index is significant at 0.05
∘ Transit is not significant
∘ Residential is significant at 0.10

Model residuals were tested for normality. We ran final iteration of the model with the above significant independent variables with lag. Model residuals are normal.

Data sources for sunlight included the COVID-19 Coronavirus Pandemic, Worldometer, (accessed on 5 May 2020 at https://www.worldometers.info/coronavirus/), Selected cities by sunshine duration, Wikipedia (accessed on 5 May 2020 at https://en.wikipedia.org/wiki/List_of_cities_by_sunshine_duration, and Selected cities by average temperature, Wikipedia (accessed on 5 May 2020 at https://en.wikipedia.org/wiki/List_of_cities_by_average_temperature). We used Pearson product-moment correlation and scatterplot visualization.

### Average daily ridership

of some cities was correlated with counts of cases and deaths where the metro was located using the Google Mobility Cross-Correlation dashboard. Transit stations had a positive correlation at +25 days lag with death count on the following locations, significant at p < 0.05 for France (0.26), New York State (0.367), and Italy (0.406). Demographics, daily ridership levels, and some national economic scores were used in correlation analysis. The plots may be viewed at https://peritusservices-my.sharepoint.com/:p:/g/personal/adrian_peritus-services_com/EU8Skd0VqvdNmgSZ_Q-x-YABmc3T2WMcACAuW_esvUU3gg?e=3asFda. Correlation coefficients for mobility data were derived from cross-correlation analysis done on each variable of the Google mobility dataset with the COVID cases and deaths and can be found at https://public.tableau.com/profile/foxyreign#!/vizhome/GlobalMobilityReport_15917747268540/GoogleMobilityReportDeaths-A4size.

The technical specifications and software used to calculate all regression models are available at https://peritusservices-my.sharepoint.com/:w:/g/personal/adrian_peritus-services_com/EQGtrtuLajBIoqxUacfwVCUB4XEPH31thzXCycV6eLQA_A?e=IZuYlr. Model selection was either based on adjusted R^2^ or f-statistic p-value using the typical Gaussian linear regression that assumes normal distribution. Log-log normal distribution was used to compute the change in deaths per million with percentage of obesity. Further information is available at https://peritusservices-my.sharepoint.com/:i:/g/personal/adrian_peritus-services_com/Eaj5eFSTyfBKtPV-YfWPAJIBLXA3FEqDgWUFQZbi3tW3Zg?e=y97F3M.

The demand for air conditioning of certain countries from 2018 and 2019 was correlated with cases and deaths. See screenshot: https://peritusservices-my.sharepoint.com/:i:/g/personal/adrian_peritus-services_com/EQZr7N-YIlpDhVogS24e0VQBf0ZsqlyjCjZPkzTqHJR3Cw?e=agsm4s. Data sources included https://www.jraia.or.jp/english/World_AC_Demand.pdf and the Cumulative COVID-19 Our World in Data cases, which showed numbers of deaths and number recovered as of 24 June 2020. We used Pearson product-moment correlation.

### Obesity

We used Our World in Data, cumulative count as of 4 Jul 2020 - https://github.com/owid/covid-19-data/blob/master/public/data/owid-covid-data.csv and the Central Intelligence Agency of obesity adult prevalence rate (Country Comparison:: Obesity - Adult Prevalence Rate, 2016) - https://www.cia.gov/the-world-factbook/field/obesity-adult-prevalence-rate/country-comparison. Countries were classified by continent according to their geographic location. Total (cumulative as of 4 Jul 2020) COVID deaths per million dependent variables was transformed using natural logarithmic + 1 to eliminate infinity values. We ran a log-normal ordinary least squares regression model using effects coding (sum-to-zero contrast, setting Africa as the reference category) with the following independent variables:

∘ Obesity prevalence rate nested with the obesity per continent
∘ Population density
∘ Aged 70 older
∘ GDP per capita
∘ COVID death rate
∘ Diabetes prevalence
∘ Female smokers
∘ Male smokers
∘ Hospital beds per thousand
∘ Life expectancy

We ran normality test on the model residuals and checked generalized variance inflation factors for multicollinearity. We ran ANOVA to check the difference in the means and then ran Tukey post-hoc tests to check for pair-wise difference in means. The model coefficients were transformed back using exponents to interpret the model values using Y percent in increase/decrease = [(exp(beta coefficient – 1) * 100].

### Face Mask Data

We used the COVID Tracking Project, accessed 8 Aug 2020, at https://covidtracking.com/data. Facemask use during the COVID-19 pandemic in the United States was accessed 8 Aug 2020 using https://en.wikipedia.org/wiki/Face_masks_during_the_COVID-19_pandemic_in_the_United_States. The COVID tracking project is a time-series dataset that has different variables with daily counts for each state. This dataset was joined with the Wikipedia dataset that classifies the dates if the counts occurred during ‘pre-mask’, ‘post-mask’ or ‘no law’. We used Quassi-poisson regression, zero-inflated Poisson regression, and auto-regressive time-series regression, plus median analysis with random sampling bootstrapped and 5,000 replications of pre-mask vs post-mask and no law.

Data was transformed to fit parametric distributions prior to statistical analyses, which were performed with Student’s t-test, linear regression and post-hoc tests. Especially for ordinary least squares regression, natural logarithmic transformations were done to remediate normality violations in the standardized residuals. Interpretation on the final model of the regression and analysis of variance were adjusted because of logarithmic transformations. For correlational studies, whether continuous or count data were used, both Pearson’s and Spearman’s rank correlational tests were done according to the data used. Normality assumptions were not made as we used non-parametric correlation tests to overcome non-linearity. Most of these normality violations related to bimodal distribution, so running Pearson’s correlation tests provided a complete description of the association [12]. Also, power in significance testing were also done to ensure that for some data points that have very minimal number of samples, this was considered that the interpretation of significance is practically taken into consideration. For cross-correlation of stochastic processes proper diagnostics were done to ensure the absence of autocorrelation processes that could signal the delayed copy of itself from its own function of lag. All significance levels are set at 0.05, otherwise specifically stated.

### Patient and Public Involvement

The research questions and outcome measures were those being discussed widely on all news channels. Given that we were all potential patients, we were all involved in the design of the study in that we asked the questions that potential patients were asking in the media.

## Results

### Natural course

Data from the Project for European Mortality Monitoring in Denmark showed that the excess mortality peak was caused by Covid-19 and not lockdown. Some countries were caught off guard by a virus that is mild enough to be invisible with a changing R value, a fairly long incubation period, and a sudden acceleration in infectivity. The countries of Spain, Italy, France, Belgium, UK, Netherlands, Switzerland, Sweden, and possibly Portugal, Malta, and Greece peaked before any lockdown took place.

Given that the median delay between infection and death lies between 21 days and 25 days (mean of 5.1 days (95% Confidence Interval (CI), 4.5 to 5.8 days) incubation to symptoms [13] and 17.8 days first symptoms to death) [13], that would place the peak of the infection in week 11 (March 9 to March 15) before lockdown of people happened and after lockdown of cluster risk places had taken place. Locking down non-essential clustering places may have been sufficient to reverse the curve that started becoming aggressive around week 8 to 9. For at least 12 weeks the epidemic was moving at a very slow pace, becoming aggressive when conditions permitted.

Temperature levels. We found no correlation for temperature, even though the epidemic did not pick up significantly where temperatures were below freezing. That may also be explained by lower population density in areas with lower temperatures. We could not find any correlations with temperature outside of an observation that infectivity did not increase when temperatures were below freezing. Wang et al. found that high temperature and high humidity reduced infectivity [14], though we found correlations only with relative humidity, hours of daylight, and levels of ultraviolet radiation. We estimated the mean incubation period to be 6.4 days (95% CI: 5.6–7.7). Backer estimated the mean duration from onset of symptoms to death to be 17·8 days (95% CI: 16·9–19·2) [15]. Using Google’s mobility data, we found a correlation between daily ridership and death count with 25 days lag. We confirmed this by checking the data of the European Mortality Monitoring Project (EuroMomo) and dates and by comparing their curves with data extracted from INSEE (Institut national de la statistique et des études économiques) in France and Statistics Belgium (Statbel) which were consistent with EuroMoMo’s data. With a reasonable degree of confidence, we traced infection peak time for countries to 3 weeks before mortality peaks to evaluate how much of its natural course the disease had in different countries. Most peoples’ lockdowns happened concurrently or after the peak had and when the curve was already diminishing.

Belgium hit a plateau of 83 deaths on April 4th with consecutively 70, 75 and 86 deaths to start dropping on April 8th, placing the probable peak and turnaround infection period around March 16th. That places the turnaround right after the lockdown of public places and before individual people lockdowned.

France hit its plateau of excess mortality on April 1st which places its probable peak around March 12th and March 14th after the ban on large gatherings, just before public places locked down, and 4 days before individual peoples were locked down. This applies also to Belgium, France, Italy, and the UK. No clear trend existed for lockdown in terms of mortality. Some public places were locked down early, some awaited outcomes and were locked down late, but we saw no clear trends to outcome for mortality in terms of when the lockdown occurred, how severe the lockdowns were, and how long they lasted. Most countries that locked down early avoided a severe peak, but Greece, Portugal, Malta, and Switzerland locked down late and also had a mild epidemic.

Countries deciding to lockdown their citizens did so as they were heading towards a high peak of infection and at the time without knowledge as to where they were in the curve except for predictions from mostly faulty epidemic models. Around April 10th (in Europe) consecutive drops began to occur each day in mortality. Studies from the Far East give us some sense as to what measures may have contributed to lowering of the curve.

From the above data, it seems reasonable to assume that the epidemic followed much of its natural course in at least 17 of 21 countries mentioned. This is further confirmed by the absence of rebounds anywhere except for localized clusters in locations/populations that had not been exposed. Regions that were exposed and where the epidemic took its course benefit from a fair level of protection under spring and summer conditions with some risk of occasional clusters but a limited risk of a full national epidemic.

Looking at a different part of the world, Japan had no lockdown, performed relatively few tests (less that most European countries), has very dense cities and one of the oldest populations, yet suffered a mild epidemic that did not seem to rebound. Japan simply instructed its population very early to avoid closed places, large crowds, and unnecessary physical contact with strangers as a policy that could be applied in the long term. Could that have sufficed? Perhaps their relatively superior diet and health helped. Tadjikistan had no lockdown and 6 deaths per million compared to Kazakstan’s (with a lockdown) of 20 deaths per million. Japan’s death rates were 8 deaths per million people; United Kingdom (UK), 650; Spain, 607; Italy, 567; United States (USA), 399; Germany, 52; and Norway, 46.

### Studying excess mortality

The increase in excess deaths in weeks 11 to 20 coincided with COVID-19 (but there’s insufficient data to prove causality). The lockdown did not occur in most of these countries until week 13 and 14 (Euromomo). Data from Weeks 1 to 23 of 2020 showed that some countries were more vulnerable to respiratory human-to-human transmitted epidemics that others, depending on population density and heating technologies. Belgium, Spain, and the UK were hardest hit (2 to 3 times influenza’s z-score peak); whereas, several countries from southern, central, and northern Europe had peaks often lower than that of recent influenza. In spite of significant peaks representing a moderate excess mortality, France had a peculiar plunge in mortality after lockdown was lifted. Except for Portugal, Greece, Germany and Austria, who are prone to epidemics and successfully “skipped their turn,” countries that had high peaks for influenza epidemics in 2017 and 2018 had high Covid-19 peaks, while those who had mild influenza epidemic peaks in 2017 and 2018 had even milder Covid-19 peaks.

In Europe, during the winter of 2020, Spain, Belgium, and the UK had a very different story than that of Portugal, Greece, Denmark or Germany with lower rates of infection. This can be particularly interesting, because of the high clustering nature of Covid and low dispersion factor. Once super-spreaders and clusters come under control, the propagation rate returns below 1 and the epidemic disappears.

### Environmental factors

For environmental/weather factors, we examined 5 key variables: ultraviolet light (UV), Ozone, Humidity, Sunshine, and Temperature. We hfound that for a variety of European and Southern hemisphere cities, there is strong association between daily UV levels [7] and deaths, as well as new cases. The association is particularly strong when lethality and new cases are lagged by 10-15 days (where the correlation coefficients range between |0.2| and |0.6|) as shown in the charts below. The amounts of sun needed may vary depending upon individual skin nature and pigmentation.

For sunshine, the 4-month average of daily sunshine is correlated with death per capita, cases per capita, death count, and case count at a borderline significance level of p = 0.1. There was no correlation for temperature. Interestingly enough, the epidemic increased in the Middle East, India and Pakistan in late spring despite high UV levels. This apparent contradiction may be explained by the population of these countries increasing their use of air conditioning, avoiding sun, and spending more time in closed places, thus creating better propagation conditions and further explaining the need for sun and fresh air to reduce spread or even reaching a point of disappearance.

The South to North gradient of the epidemic curve dropped in Europe as spring arrived, combined with lower peaks in countries where their parks were open and people could access sun, further indicating a beneficial effect of sun or UV’s or light or vitamin D (Mitchell, 2020).

The final model has a very low coefficient of determination of 0.0098 (meaning that the variables included in the model only explain 0.98% of the variation with the % change of deaths but this is statistically significant at p < 0.05:

- A unit of decrease of UV Index after 7 days, on average, increases the % change in daily deaths.
- A unit of increase in residential % change from baseline after 10 days, on average, increases the % change in daily deaths.

### Ozone index levels

For Ozone, the correlation was weak, and it was discarded We found an inverse correlation between humidity levels and absolute death counts.

March average relative humidity % (−0.63, p < 0.01)

January average relative humidity % (−0.58, p < 0.05)

February average relative humidity % (−0.56, p < 0.05)

This humidity correlation may contribute to explain why coastal cities often faired better. London, Madrid, Brussels, Milan, and Paris were the hardest hit in Europe in that order, while none are coastal. The hardest hit coastal cities were Barcelona, Stockholm, and Amsterdam, which had either no lockdown or a very mild lockdown. Humidity did not appear to be a factor In Figure 1, we present the Cross-Correlation of UV index and Ozone for several cities:

**Figure 1.**
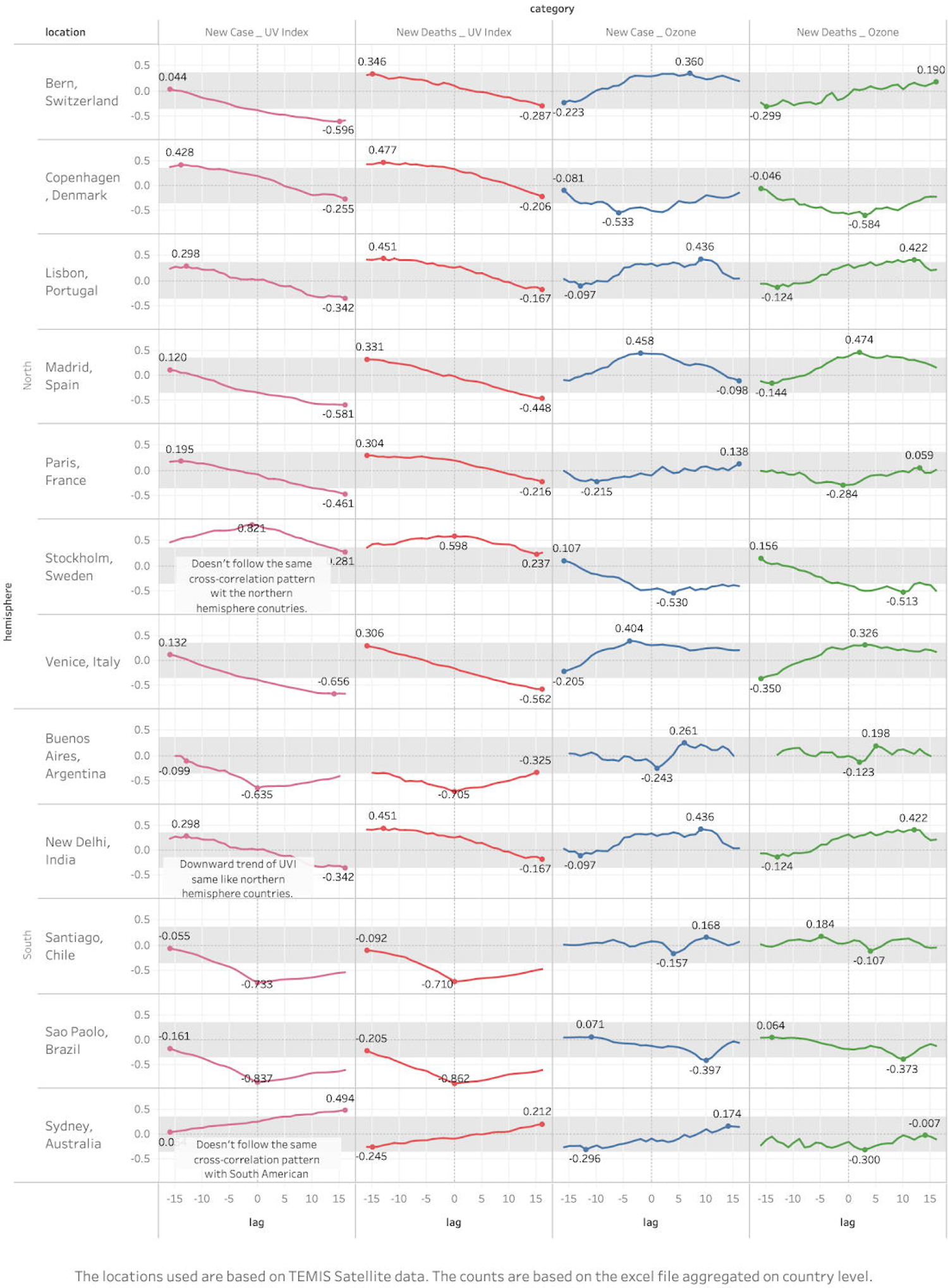
Cross-Correlation of UV index and Ozone for several cities.

### Socio-economic Factors

For social variables, we looked at two sets of data; demographic data (Density per km2, average daily ridership for cities with a metro, the Gini index of income distribution, the gross domestic product (GDP) per capita, and mobility data (changes in frequenting of residential, workplaces, parks, grocery/pharmacy, retail/recreation and transit stations).

The following correlations (and significance levels) were found.

Positive correlation with case count:

- Average daily ridership on publica transportation: 0.73, p < 0.01
- Density km2: 0.61, p < 0.05
- GINI Index: 0.57, p < 0.10 Positive correlation with death count, p < 0.10:
- Average daily ridership: 0.58
- Density km2: 0.52 Positive correlation with cases per 100k, p < 0.10:
- Density km2: 0.57

This shows that increases in ridership and higher population densities are associated with both higher infection rates and deaths and that higher income inequality (as measured by Gini) is associated with a higher number of cases. In developing countries, the epidemic seems to hit wealthier populations harder; whereas, in Europe and United States, it seems to hit disadvantaged populations harder.

These variables were statistically significant to have moderate inverse correlation with Deaths per 100k at 0.10 significant level:

- 4-month average of daily sunshine (−0.46)
- March average relative humidity % (−0.45)
- 4-month average relative humidity % (−0.42)

Figure 2 shows the correlation matrix of socio-economic variables for cities and countries aggregated together

**Figure 2.**
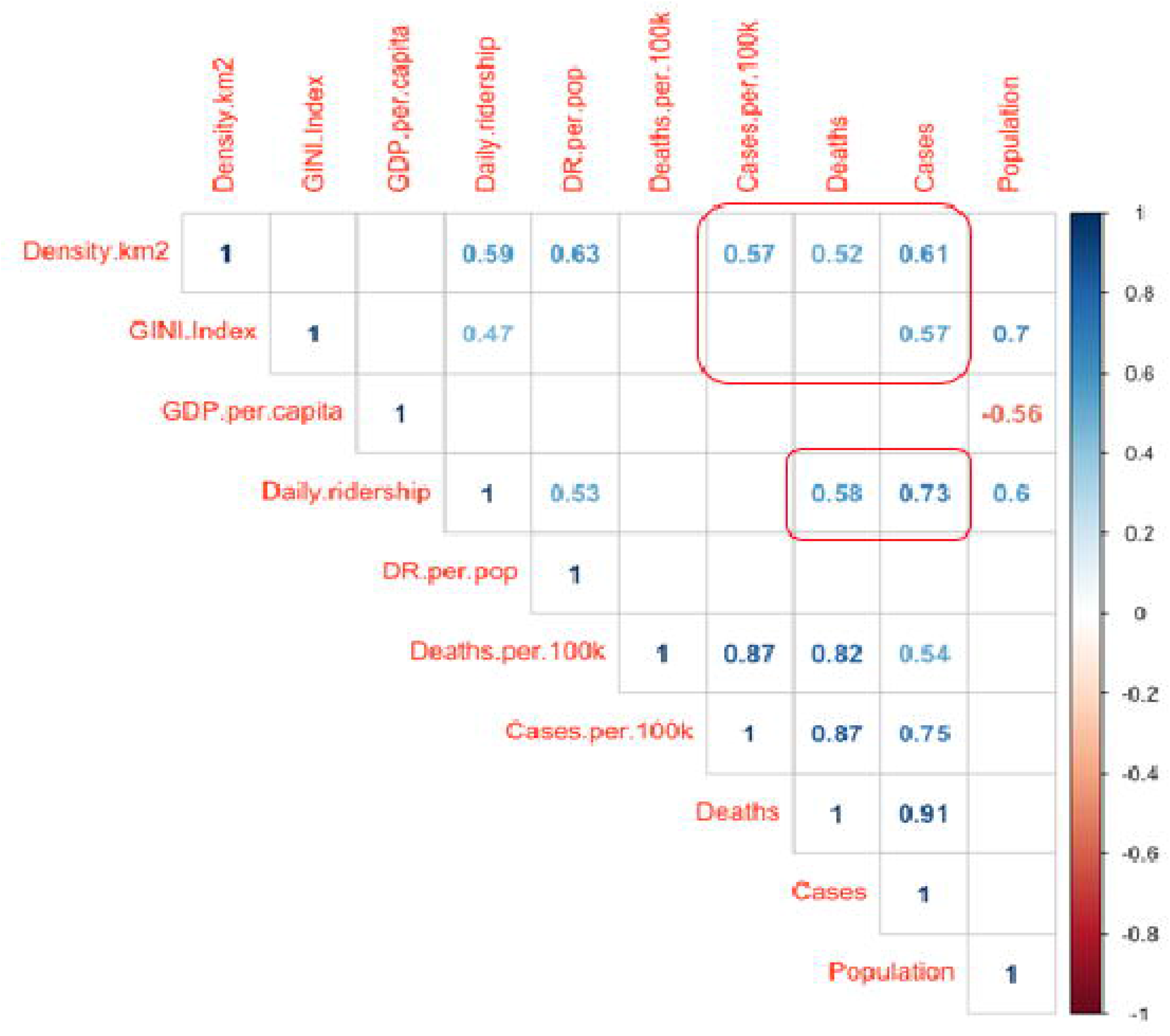
Correlation matrix of socio-economic variables for cities and countries aggregated together.

### Mobility data

Using Google mobility data, we have found that there is a statistically significant correlation in most countries at +25 lag between the presence of transit stations and deaths from COVID-19. This pattern also applies to workplaces in countries with mild or no lockdown. Metro/Subway ridership is often an indication of office concentrations in modern buildings with shut windows, possibly recycled air or HVAC where clusters may form. Both variables may correlate. Did metro mass transportation contribute to the high infection rate or was it office buildings or a combination (as there’s high correlation between daily death count and cases count). Significance was set at 0.05, df = 25, so that the correlation coefficient should be > |0.22|. To a lesser extent a similar pattern applies to retail and recreation. This is altered in the sense that most recreation and retail had been closed in almost all countries and restricted in Sweden. All five hardest hit cities were thickly populated and had a dense subway system (London, New York, Madrid, Brussels, Milan, Paris). Figure 3 shows the cross-correlation coefficients of Google mobility variables with Covid-19 death counters per location.

**Figure 3.**
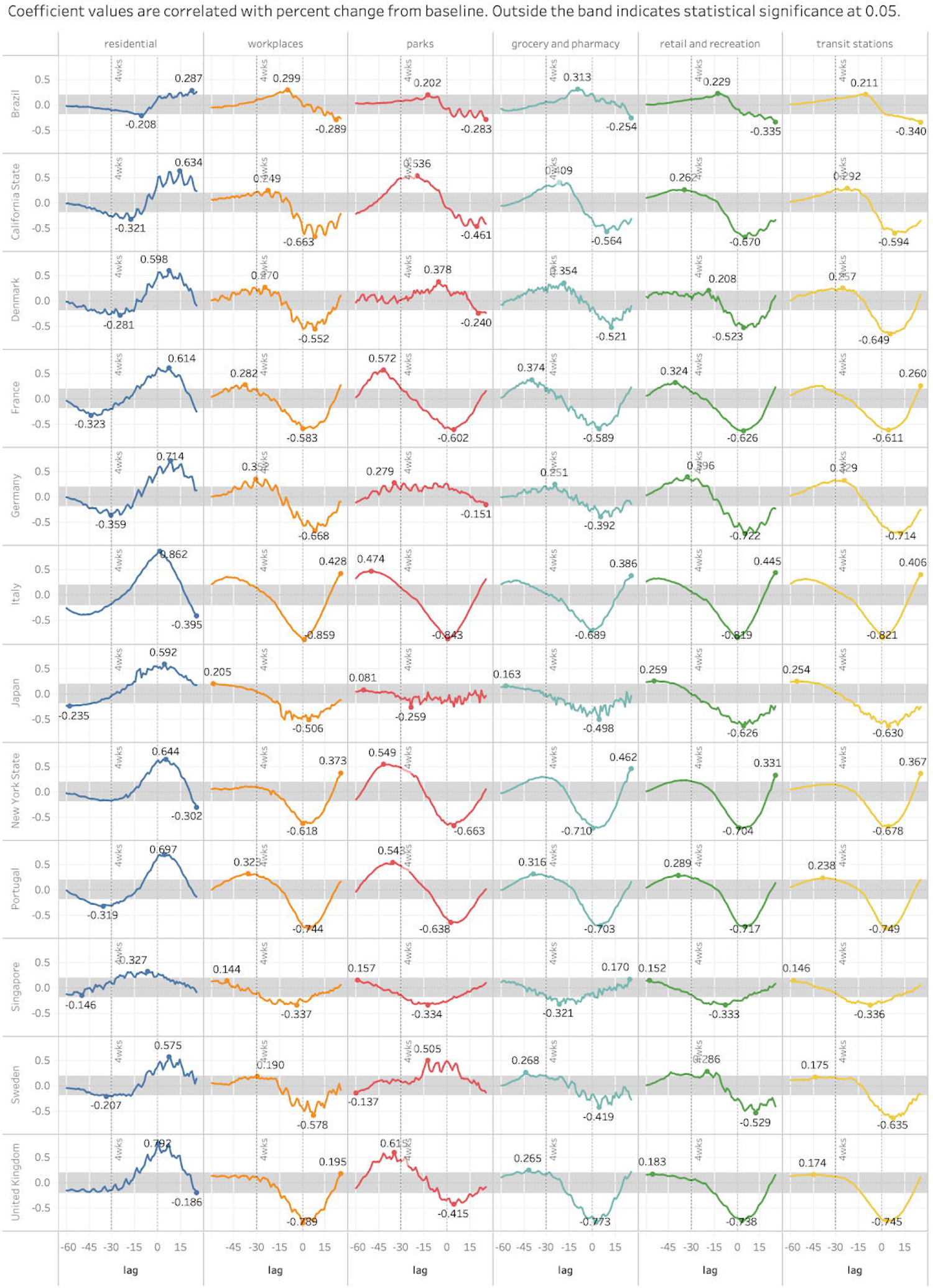
Cross-correlation coefficients of Google mobility variables with Covid-19 death counters per location.

### Diet, lifestyle, and obesity

Early in the epidemic, it appeared that co-morbidities played a role in the Covid-19 impact, including diabetes and obesity, advanced age (where co-morbidity is much more common). To quantify the impact of lifestyle variables on Covid-19 mortality, we turned to “Our World In Data,” which provides daily, collated, open source data for Covid-19 mortality (using the European Centre for Disease Prevention and Control, the World Health Organization and Johns Hopkins University). We used the latest cumulative natural log transformed total death per million and regressed it against the 2016 compilation of the adult prevalence rate of obesity [10]

In our first run, in which we did a worldwide regression, we found that the obesity adult prevalence rate (which is the proportion of obese people in the population per country) was a statistically significant predictor in determining the Total Deaths Per Million. For each unit of increase in obesity %, the average total death per million increased by 8.76% on a global average (with a highly significant p-value). However, as the epidemic was at different stages in different continents, we did the regression by continent. This gave us the following results: Africa (coefficient = 6.48; p = 0.0078), South America (coefficient = 10.88, p = 0.000048), North America (coefficient = 9.07, p = 0.000049), Europe (coefficient = 12.89, p = 0.000041), Asia (coefficient = 8.51, p = 0.00057), Oceania (coefficient = −1.42, p = 0.6193).

For each continent, we found:

- Oceania: each unit increase in Oceania obesity% means 1.42 % increase in Oceania’s total deaths per million (note there are only 3 countries here: Fiji, NZ and Australia, also not significant)
- South America: each unit increase in South America obesity % means 10.88% increase in
- South America total deaths per million
- North America: each unit increase in North America obesity % means 9.07% increase in North America s total deaths per million
- Europe: each unit increase in Europe obesity % means 12.89% increase in Europe total deaths per million
- Asia: each unit increase in Asia obesity % means 8.51% increase in Asia’s total deaths per million
- Africa: each unit increase in Africa obesity % means 6.48 % increase in Africa’s total deaths per million.

Oceana’s countries closed their borders very early. In the presence of obesity, other variables (diabetes prevalence, smoking, age 70 years or older, life expectancy, population density) were insignificant, further demonstrating the importance of obesity as a significant indicator in the epidemic course. Japan, the country with the oldest population in the world, with very high population density, with no lockdown, few restrictions and low testing outperformed by most other countries, has the lowest obesity rate in the world at 4.3% and had minimal deaths. Others have confirmed these observations [16, 17]

### Air Conditioning (AC) or HVAC

Most modern AC units are in fact HVAC. Our analysis of available AC demand showed a moderate correlation between commercial demand as per the Japan Refrigeration and Air Conditioning Association (JRAIA) 2018 data and death from Covid-19 until June 25th at significance level of p < 0.1 (R = 0.65) using 15 European countries which are at the end of their epidemic wave and for which data was available. This was run on a limited sample size (n=12). At the 0.1 significance cutoff, the probability to reject a false negative is 25%; the sample size is sufficient at the expected correlation coefficient of 0.65. Figure 4 shows the correlation coefficients related to air conditioning.

**Figure 4.**
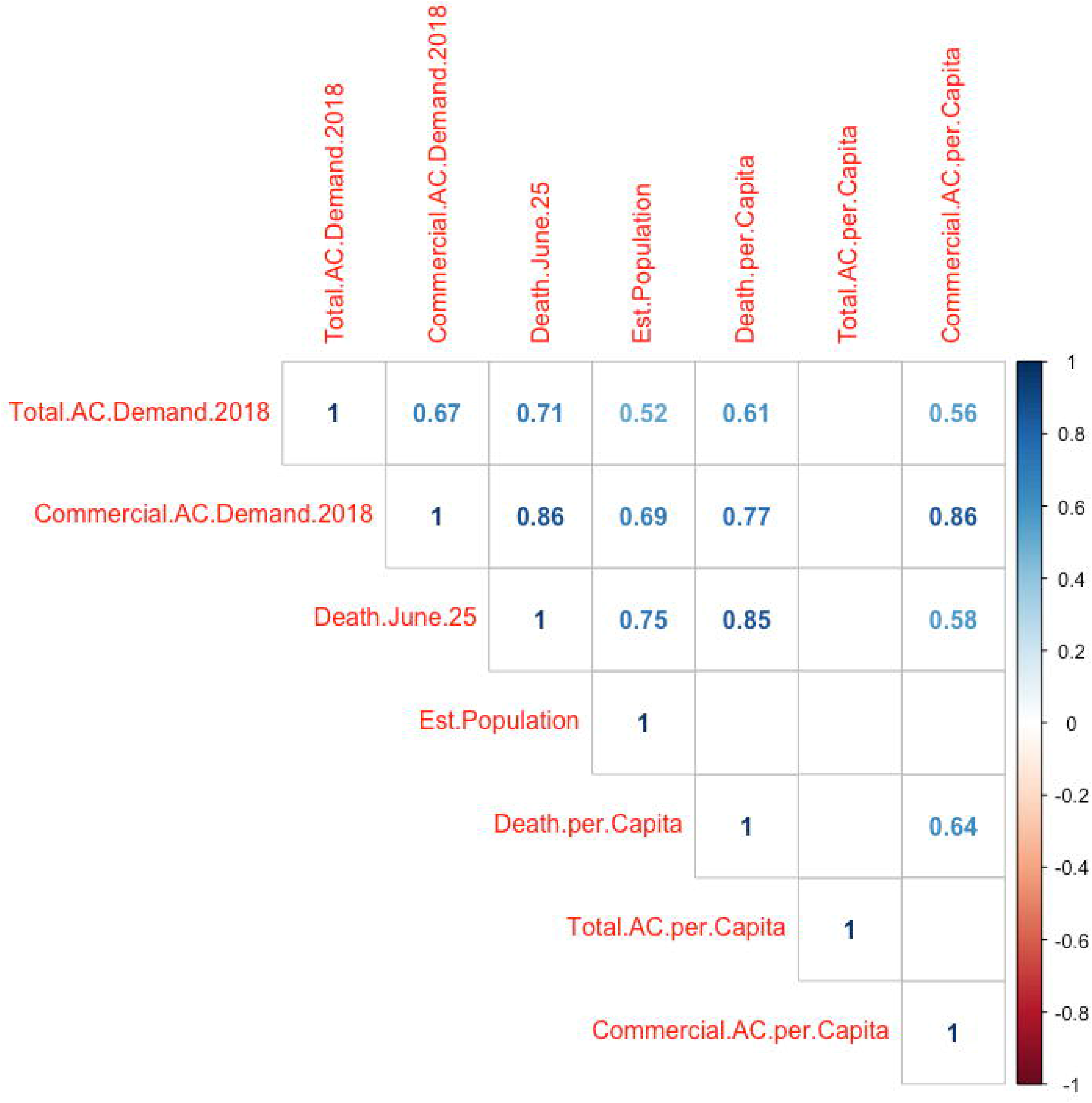
Correlation coefficients related to air conditioning.

The absolute death count as of June 25 is correlated with 2018 commercial AC demand at 0.86; 2018 total AC demand at 0.71; and commercial AC per capita at 0.58. Death per capita was associated with 2018 commercial AC demand at 0.77; commercial AC per capita at 0.64; and 2018 Total AC demand 2018 at 0.61 With regard to influenza spread, improvement of ventilation in high density public spaces could be an important and relatively easy-to-implement strategy supplementing vaccination, corresponding to a vaccination coverage of 60– 70% for an efficacy of 40%, 50–60% for 60% efficacy, and 40–50% for 80% efficacy. In the aerosol model, consistently improved ventilation beats vaccination even with full coverage if efficacies are low. It can be done fast and it works for aerosols, known droplets, and yet unknown viruses [18]. All five hardest hit cities had a high concentration of modern office buildings and hotels with shared and/or a culture of centralized HVAC in office buildings, malls, homes (London, New York, Madrid, Brussels, Milan). For most buildings, the easiest way to deliver outside air directly across the building envelope is to open a window. Window ventilation not only bypasses ductwork but increases outside air fraction and increases total air change rate as well (79). Administrators and building operators should discuss a plan for increasing perimeter, and specifically window, ventilation when outdoor temperatures are adequate for this practice [19].

### Blood type

We found no relation with blood type.

### Masks

None of the generalized regression models for predictions based upon the use of facemasks were valid. The time-series AR did not finish due to RAM constraints.

Asian countries easily wore masks and were spared. Denmark, Finland, and Norway skipped masks and were spared. African countries tried to wear masks and took them off and were spared. Open air music festivals and demonstrations on June 21st with or without masks did not show an increase in cases or the appearance of clusters within 14 days.

Given a median delay between infection and symptoms of 5.1 days, depending upon testing policies, the effects of a mandatory mask wearing policy should appear within 5 days and become definitive within 14 days. The shape of the curve for California did not change despite a mandatory masks policy in the entire state on 18 June, 25 days after they went to lockdown (Figure 5) (Data from https://covidtracking.com/data/download). Similar findings occurred in Texas and in France. This raises questions about the degree of compliance with mask wearing and the effects of different types of masks worn given a mandatory mask-wearing policy.

**Figure 5.**
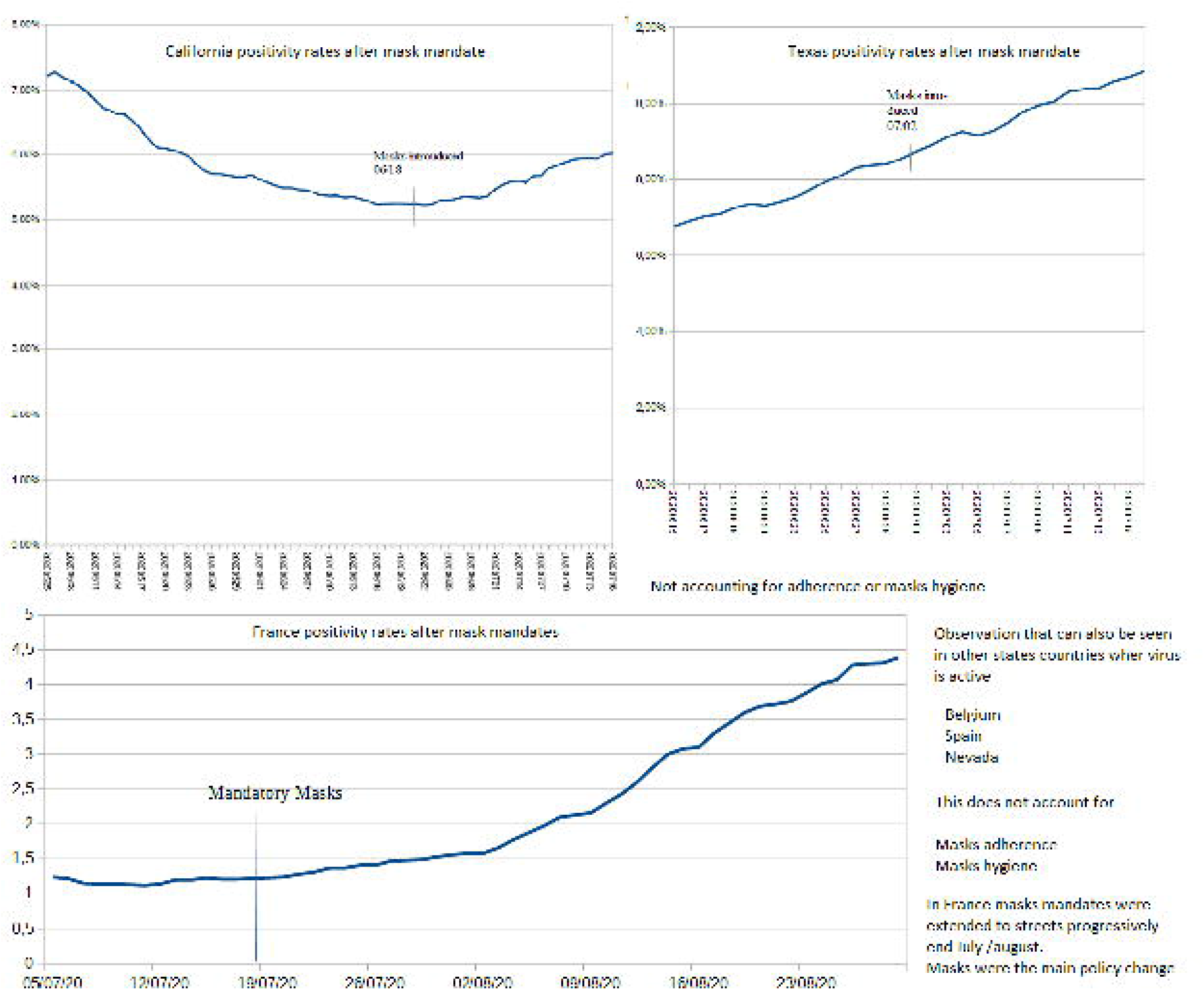
Effects of mandatory mask wearing policies on the shape of the curve for Covid positivity rates for California, Texas, and France.

## Conclusion

From the multiple factors impacting the epidemic that we analyzed, and, comparing multiple countries across the world at a macro level on the one side and verifying occurrence at micro level through studies review or cluster analysis, 2 factors stand out as to predicting severity: (1) UV/Vitamin D levels and (2) the health of the population and especially obesity. At the time of writing this paper, we found no country with an obesity level under 8% having a severe epidemic. We also found that countries in which the population benefited from sun exposure or vitamin D supplementation and spent time outside, fared well.

A few factors stand out as to propagation: (1) Commercial HVAC, (2) Density and poorly aerated gatherings, (3) Relative humidity; (4) Level of UV, (5) Timely policies of limiting gatherings in closed clustered places until aeration is improved, and (6) reducing daily Metro ridership.

Multiple other factors explained little of the variance including population lockdowns, mask-wearing policies, blood types, and ozone. However, mask policies do not tell us the degree to which people complied with mask policies or the effectiveness of types of masks worn. Masks may have a mitigating role in situations of high density with HVAC or travel on poorly ventilated public transportation. Dispersing urban dwellers into the countryside and away from high occupancy buildings with HVAC could reduce transmission. Contact tracing was not analyzed as very few countries applied it for long enough. Excess mortality observed is within ranges of severe past influenza epidemics 2016/2017 or 1999/2000 and lower than older influenza epidemics of the 1940s or 1970s.

Treatments or vaccines may protect the fraction of the population not suffering from severe comorbidities. Prevention measures such as reviewing aeration systems, enhancing diet and exercise, and ensuring adequate levels of vitamin-D may turn out to be efficient in providing protection against COVID, possibly influenza, and other epidemics, not to mention increasing the efficiency and wellbeing of populations.

The use of large databases with aggregate data has its limitations in that individual variation can be obscured.

## Data Availability

All data is available for sharing.

http://www.propagmath.org

## Contributorship Statement

Drs. Sellier and Bricaire conceived the project and assembled the team to conduct the project. Identification of appropriate databases and statistical analysis of data within those databases was conducted by Drs. Cuyugan, Barac, Parvaiz, Bin Jamil, Iqbal, Vally, Koliali, and Sellier. Interpretation of the findings involved all the authors. Dr. Sellier and Dr. Mehl-Madrona wrote the article.

## Competing Interests

### Data Sharing

The URLs for all datasets used are provided in the text of the article and are listed as references 1 through 11.

## Abbreviations used

SARSCOV-2: the most recent version of the coronavirus leading to the current pandemic
HVAC: heating, ventilation, and air conditioning
CI: Confidence Interval
INSEE: Institut national de la statistique et des études économiques
**E**UROMOMO: European Mortality Monitoring Project
Statbel: Statistics Belgium
UK: United Kingdom
USA: United States of America
UV: ultraviolet light
GDP: gross domestic product
AC: air conditioning
JRAIA: Japan Refrigeration and Air Conditioning Association
P: probability

## Notes

The authors have no conflict of interest to report.

### Competing Interest Statement

The authors have declared no competing interest.

### Funding Statement

No external fundng

### Summary of Updates

We have greatly expanded the methods section of this paper.

